# Overcoming data gaps in life course epidemiology by matching across cohorts

**DOI:** 10.1101/2020.07.21.20158857

**Authors:** Katrina L. Kezios, Scott Zimmerman, Kara Rudolph, Sebastian Calonico, Adina Zeki Al-Hazzouri, M. Maria Glymour

## Abstract

Lifecourse epidemiology is hampered by the absence of studies with exposures and outcomes measured at different life stages. We describe when the effect of an exposure (A) on an outcome (Y) in a target population is identifiable in a combined (“synthetic”) cohort created by pooling an early-life cohort including exposure measures with a late-life cohort including outcome measures. We enumerate the causal assumptions needed for unbiased effect estimation in the synthetic cohort and illustrate by simulating target populations under four causal models. From each population, we drew hypothetical early- and late-life cohorts and created a synthetic cohort by matching individuals from the two cohorts based on mediators and/or confounders. We compared bias when estimating the effect of A on Y in the synthetic cohort, varying which matching variables were available, the match ratio, and the distance matching criterion. When the set of matching variables includes all variables d-connecting exposure and outcome (i.e., variables blocking all back and front door pathways), the synthetic cohort yields unbiased effect estimates. Methods based on merging cohorts provide opportunities to hasten the evaluation of early- and mid-life determinants of late life health, but rely on available measures of both confounders and mediators.

## Introduction

Life course epidemiology posits that risk of disease at any given age may be influenced by experiences much earlier in life, and even prenatal exposures.^1^ Understanding these early- and mid-life determinants of late-life disease is often essential for primary prevention. Ideally, to answer life course research questions we would use data from large and diverse birth cohorts with complete follow-up over the full life span that include frequent detailed social and health assessments over time. Unfortunately, few birth cohort studies are available in the United States or many other countries,^2-5^ and no single study can comprehensively measure all health outcomes potentially of scientific interest. In the absence of the ideal study, life course epidemiology research could instead draw information from multiple studies with longitudinal information on individuals at different life stages. Combining such cohorts could open new opportunities to evaluate novel effects of early/mid-life exposures on later life health.

A small number of initiatives have pooled studies from across the life course to create synthetic cohorts.^6-20^ Within this literature, limited attention has been paid to the causal conditions under which unbiased effect estimation from synthetic cohorts is achievable.^21^ Recent work on the “data fusion” problem provides a formal causal inference framework for pooling heterogeneous data sets,^22-25^ focused on transporting models from one population to another. However, data fusion methods have seen limited application in epidemiology,^23,25-27^ especially life course epidemiology.

In this article, we build on methods for data pooling and transport developed within causal frameworks^22,25,28,29^ to evaluate an approach to combining cohorts across the life course based on the assumption that multiple cohorts are drawn from a common target population at different ages (throughout, we note how this assumption can be relaxed). Conceptually, if two cohorts are drawn from the same population, the later life cohort would represent what happens to the earlier life cohort as it ages. Under this assumption, we evaluate a flexible and intuitive method for creating a synthetic life course cohort using one-to-many matching to combine individuals from a later-life cohort with their likely counterparts from an early-life cohort, where matching is based on a carefully selected set of covariates measured in both cohorts. We demonstrate using simulations the causal structures and conditions under which such a synthetic cohort can be used to estimate causal effects of exposures measured in the earlier-life cohort on outcomes measured in the later-life cohort and illustrate the bias when the assumptions are not met. For clarity, we use the example of estimating the effect of elevated early-life blood pressure on later-life dementia risk.

## Methods

### Overview

Our study is motivated by the question: when is the average treatment effect (ATE) of an exposure on an outcome identifiable in a combined (“synthetic”) cohort in which measures of the exposure and outcome are not available in the same data source? Informed by the methodological literature (e.g., data fusion, transportability, causal inference),^22,23,28,30-38^ to answer this question we first enumerate the conditions required for pooling cohorts and the causal assumptions necessary for identification in the synthetic cohort. We then conduct a simulation study to demonstrate these conditions under multiple example causal scenarios, using a one-to-many matching approach to combine cohorts.

#### (1) Notation and causal models

We define 4 nonparametric structural causal models representing the relationships between 5 variables in a target population, and formally encoded by causal directed acyclic graphs (DAGs, **Figure 1**).^39^ These variables include: the early-life exposure of interest A [*e*.*g*., early life elevated blood pressure], the late-life outcome of interest, Y [*e*.*g*., dementia risk score], variables M_1_ and M_2_ which mediate the relationship between A and Y [*e*.*g*, midlife life elevated blood pressure and midlife elevated cholesterol], and a variable C that confounds the relationship between A and Y [*e*.*g*., childhood socioeconomic status (SES)]. **Table 1** defines the data generating rule for each variable under each causal model in **Figure 1**. In brief, in DAG 1, M_1_ fully mediates the effect of A on Y. In DAG 2, M_1_ partially mediates the effect of A on Y but there is also a direct effect from A to Y. DAG 3 builds on DAG 2, adding a node (M_2_) on an additional direct path from A to Y; in one case M_1_ also causes M_2_ (DAG 3a) and in another M_2_ causes M_1_ (DAG 3b). Finally, DAG 4 modifies DAG 1 by adding a node C which creates a backdoor path between A and Y. Throughout, we assume all covariates are continuous variables (although we note that this assumption can be relaxed).

**Table 1.** Parameter inputs for simulation scenarios^1^

| DAG | Scenario<sup>1</sup> | Data generating rules | Parameter inputs |
| --- | --- | --- | --- |
| 1 | | $f_a: A = \alpha_1 U_A$<br>$f_{m1}: M_1 = \beta_1 A + \beta_2 U_{M1}$<br>$f_y: Y = \gamma_1 M_1 + \gamma_2 U_Y$ | $\alpha_1 = 0.10$<br>$\beta_1 = 0.50 ; \beta_2 = 0.10$<br>$\gamma_1 = 0.60 ; \gamma_2 = 0.10$ |
| 2 | | $f_a: A = \alpha_1 U_A$<br>$f_{m1}: M_1 = \beta_1 A + \beta_2 U_{M1}$<br>$f_y: Y = \gamma_1 A + \gamma_2 M_1 + \gamma_3 U_Y$ | $\alpha_1 = 0.10$<br>$\beta_1 = 0.50 ; \beta_2 = 0.10$<br>$\gamma_1 = 0.10 ; \gamma_2 = 0.40 ; \gamma_3 = 0.10$ |
| 3a | A, B | $f_a: A = \alpha_1 U_A$<br>$f_{m1}: M_1 = \beta_1 A + \beta_2 U_{M1}$<br>$f_{m2}: M_2 = \theta_1 A + \theta_2 M_1 + \theta_3 U_{M2}$<br>$f_y: Y = \gamma_1 M_1 + \gamma_2 M_2 + \gamma_3 U_Y$ | $\alpha_1 = 0.10$<br>$\beta_1 = 0.30 ; \beta_2 = 0.10$<br>$\theta_1 = 0.40 ; \theta_2 = 0.30 ; \theta_3 = 0.10$<br>$\gamma_1 = 0.38 ; \gamma_2 = 0.38 ; \gamma_3 = 0.10$ |
| 3b | A, B | $f_a: A = \alpha_1 U_A$<br>$f_{m1}: M_1 = \beta_1 A + \beta_2 M_2 + \beta_3 U_{M1}$<br>$f_{m2}: M_2 = \theta_1 A + \theta_2 U_{M2}$<br>$f_y: Y = \gamma_1 M_1 + \gamma_2 M_2 + \gamma_3 U_Y$ | $\alpha_1 = 0.10$<br>$\beta_1 = 0.40 ; \beta_2 = 0.30 ; \beta_3 = 0.10$<br>$\theta_1 = 0.30 ; \theta_2 = 0.10$<br>$\gamma_1 = 0.38 ; \gamma_2 = 0.38 ; \gamma_3 = 0.10$ |
| 4 | A, D | $f_c: C = \delta_1 U_C$<br>$f_a: A = \alpha_1 C + \alpha_2 U_A$<br>$f_{m1}: M = \beta_1 A + \beta_2 U_M$<br>$f_y: Y = \gamma_1 C + \gamma_2 M + \gamma_3 U_Y$ | $\delta_1 = 0.05$<br>$\alpha_1 = 0.65 ; \alpha_2 = 0.05$<br>$\beta_1 = 0.60 ; \beta_2 = 0.05$<br>$\gamma_1 = 0.65 ; \gamma_2 = 0.50 ; \gamma_3 = 0.05$ |
| | B, E | $f_c: C = \delta_1 U_C$<br>$f_a: A = \alpha_1 C + \alpha_2 U_A$<br>$f_{m1}: M = \beta_1 A + \beta_2 U_M$<br>$f_y: Y = \gamma_1 C + \gamma_2 M + \gamma_3 U_Y$ | $\delta_1 = 0.05$<br>$\alpha_1 = 0.65 ; \alpha_2 = 0.05$<br>$\beta_1 = 0.60 ; \beta_2 = 0.05$<br>$\gamma_1 = 0.1 ; \gamma_2 = 0.50 ; \gamma_3 = 0.05$ |
| | C, F | $f_c: C = \delta_1 U_C$<br>$f_a: A = \alpha_1 C + \alpha_2 U_A$<br>$f_{m1}: M = \beta_1 A + \beta_2 U_M$<br>$f_y: Y = \gamma_1 C + \gamma_2 M + \gamma_3 U_Y$ | $\delta_1 = 0.05$<br>$\alpha_1 = 0.15 ; \alpha_2 = 0.05$<br>$\beta_1 = 0.60 ; \beta_2 = 0.05$<br>$\gamma_1 = 0.65 ; \gamma_2 = 0.50 ; \gamma_3 = 0.05$ |

<sup>1</sup>Across all scenarios, we assume all exogenous U terms ( $U_A, U_M, U_{M1}, U_{M2}, U_C, U_Y$ ) are independent with a standard normal distribution,

**Figure 1.**
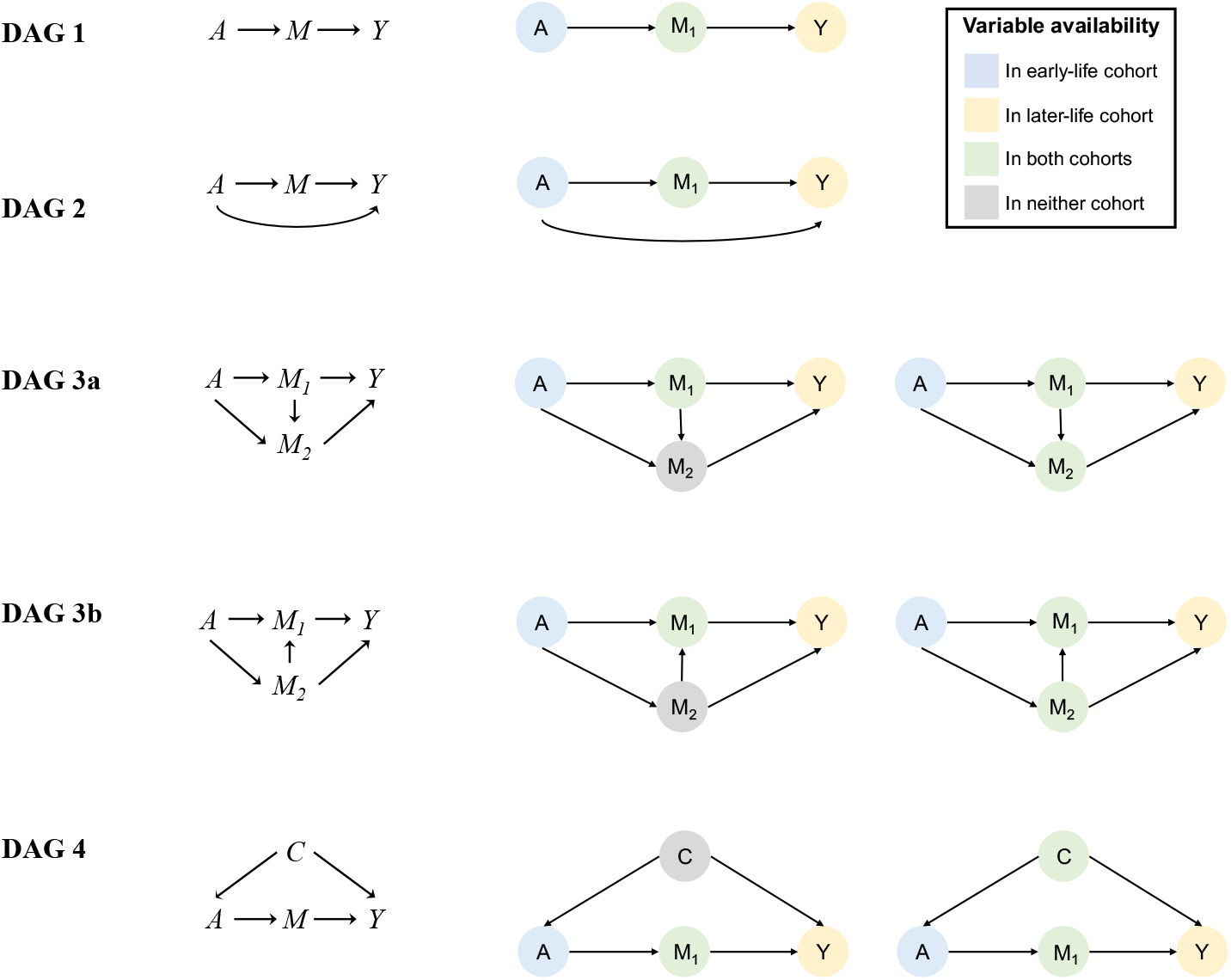
Causal scenarios considered Figure 1 shows the structural causal models depicted as directed acyclic graphs considered in the simulation study. As a visual aid to link the causal scenarios with the variables available in the observed data across different simulation scenarios, to the right these DAGs are redrawn and color-coded to show in which cohort (early-life, late-life, or both) variables are measured. Variables in the DAGs are blue if only measured in the early-life cohort (i.e., A), yellow if only in the late-life cohort (i.e., Y), or green if measured in both (e.g., M_1_). Variables that are part of causal structure underlying the data but are not measured in either cohort are gray (e.g., M_2_). For DAGs 1 and 2, only one simulation scenario is considered where the merging variable is available in both cohorts. For DAGs 3a, 3b, and 4 multiple simulation scenarios are conducted based on variable availability (e.g., for DAG 3a we conduct a scenario where M_2_ is unavailable in both cohorts and when it is available in both cohorts). All simulation scenarios are summarized in Table 2.

#### (2) Conditions for Combining Cohorts and Identification

In general, to allow unbiased inferences about a common target population from the combined, synthetic life course cohort, the relationships between all the variables in the target population must be reproduced in the synthetic cohort upon merging of the early- and late-life cohorts. We represent cohort membership as R=0 (early life cohort) or R=1 (late life cohort) in equations, which could analogously be represented in the DAGs as an additional R node. However, throughout we assume that both cohorts are simple random samples from the target population and therefore unrelated to any other variable shown in the DAG. This means that on the DAGs in Figure 1, R would be represented as a stand-alone node with no entering or exiting arrows; because of this, we omit the R node from the DAGs.

**Table 2.**
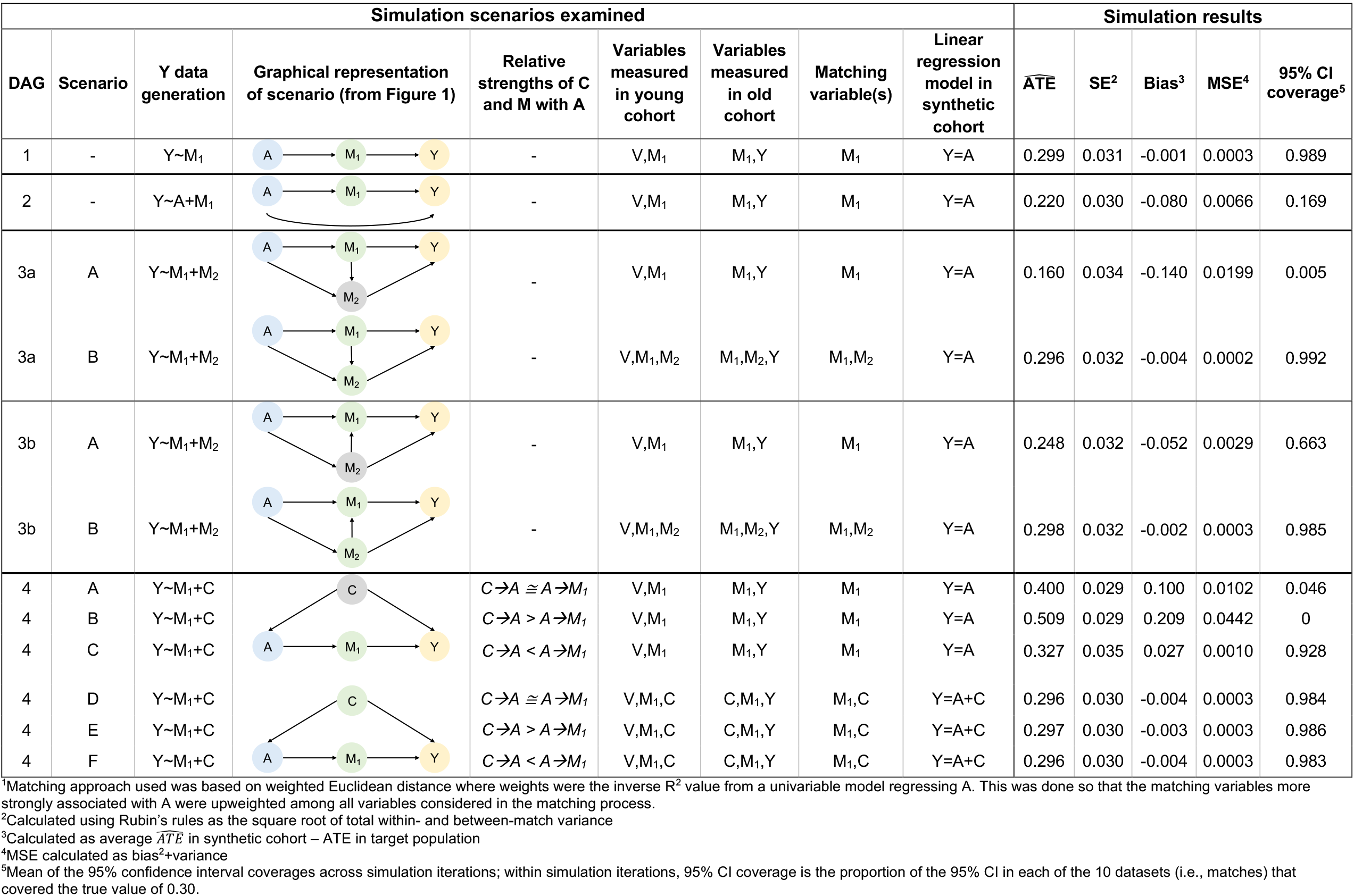
Details of the twelve simulation scenarios examined and their results in the synthetic cohort (N=2,500) created using 1:10 matching^1^ across 1,000 simulation iterations

| Simulation scenarios examined |  |  |  |  |  |  |  |  | Simulation results |  |  |  |  |
| --- | --- | --- | --- | --- | --- | --- | --- | --- | --- | --- | --- | --- | --- |
| DAG | Scenario | Y data generation | Graphical representation of scenario (from Figure 1) | Relative strengths of C and M with A | Variables measured in young cohort | Variables measured in old cohort | Matching variable(s) | Linear regression model in synthetic cohort | $\widehat{ATE}$ | SE<sup>2</sup> | Bias<sup>3</sup> | MSE<sup>4</sup> | 95% CI coverage<sup>5</sup> |
| 1 | - | $Y \sim M_1$ | | - | V, M<sub>1</sub> | M<sub>1</sub>, Y | M<sub>1</sub> | $Y=A$ | 0.299 | 0.031 | -0.001 | 0.0003 | 0.989 |
| 2 | - | $Y \sim A + M_1$ | | - | V, M<sub>1</sub> | M<sub>1</sub>, Y | M<sub>1</sub> | $Y=A$ | 0.220 | 0.030 | -0.080 | 0.0066 | 0.169 |
| 3a | A | $Y \sim M_1 + M_2$ | | - | V, M<sub>1</sub> | M<sub>1</sub>, Y | M<sub>1</sub> | $Y=A$ | 0.160 | 0.034 | -0.140 | 0.0199 | 0.005 |
| 3a | B | $Y \sim M_1 + M_2$ | | - | V, M<sub>1</sub>, M<sub>2</sub> | M<sub>1</sub>, M<sub>2</sub>, Y | M<sub>1</sub>, M<sub>2</sub> | $Y=A$ | 0.296 | 0.032 | -0.004 | 0.0002 | 0.992 |
| 3b | A | $Y \sim M_1 + M_2$ | | - | V, M<sub>1</sub> | M<sub>1</sub>, Y | M<sub>1</sub> | $Y=A$ | 0.248 | 0.032 | -0.052 | 0.0029 | 0.663 |
| 3b | B | $Y \sim M_1 + M_2$ | | - | V, M<sub>1</sub>, M<sub>2</sub> | M<sub>1</sub>, M<sub>2</sub>, Y | M<sub>1</sub>, M<sub>2</sub> | $Y=A$ | 0.298 | 0.032 | -0.002 | 0.0003 | 0.985 |
| 4 | A | $Y \sim M_1 + C$ | | $C \rightarrow A \cong A \rightarrow M_1$ | V, M<sub>1</sub> | M<sub>1</sub>, Y | M<sub>1</sub> | $Y=A$ | 0.400 | 0.029 | 0.100 | 0.0102 | 0.046 |
| 4 | B | $Y \sim M_1 + C$ | | $C \rightarrow A > A \rightarrow M_1$ | V, M<sub>1</sub> | M<sub>1</sub>, Y | M<sub>1</sub> | $Y=A$ | 0.509 | 0.029 | 0.209 | 0.0442 | 0 |
| 4 | C | $Y \sim M_1 + C$ | | $C \rightarrow A < A \rightarrow M_1$ | V, M<sub>1</sub> | M<sub>1</sub>, Y | M<sub>1</sub> | $Y=A$ | 0.327 | 0.035 | 0.027 | 0.0010 | 0.928 |
| 4 | D | $Y \sim M_1 + C$ | | $C \rightarrow A \cong A \rightarrow M_1$ | V, M<sub>1</sub>, C | C, M<sub>1</sub>, Y | M<sub>1</sub>, C | $Y=A+C$ | 0.296 | 0.030 | -0.004 | 0.0003 | 0.984 |
| 4 | E | $Y \sim M_1 + C$ | | $C \rightarrow A > A \rightarrow M_1$ | V, M<sub>1</sub>, C | C, M<sub>1</sub>, Y | M<sub>1</sub>, C | $Y=A+C$ | 0.297 | 0.030 | -0.003 | 0.0003 | 0.986 |
| 4 | F | $Y \sim M_1 + C$ | | $C \rightarrow A < A \rightarrow M_1$ | V, M<sub>1</sub>, C | C, M<sub>1</sub>, Y | M<sub>1</sub>, C | $Y=A+C$ | 0.296 | 0.030 | -0.004 | 0.0003 | 0.983 |

<sup>1</sup>Matching approach used was based on weighted Euclidean distance where weights were the inverse $R^2$ value from a univariable model regressing A. This was done so that the matching variables more strongly associated with A were upweighted among all variables considered in the matching process.
<sup>2</sup>Calculated using Rubin's rules as the square root of total within- and between-match variance
<sup>3</sup>Calculated as average $\widehat{ATE}$ in synthetic cohort – ATE in target population
<sup>4</sup>MSE calculated as bias<sup>2</sup>+variance
<sup>5</sup>Mean of the 95% confidence interval coverages across simulation iterations; within simulation iterations, 95% CI coverage is the proportion of the 95% CI in each of the 10 datasets (i.e., matches) that covered the true value of 0.30.

We focus on identifying a causal estimand, such as the average treatment effect (ATE) of a continuous exposure A on a continuous outcome Y in the target population, defined as *E*[*Y*_*a*_ − *Y*_*a*_*] where *Y*_*a*_ and *Y*_*a*_* represent an individual’s counterfactual outcomes under exposure values A=a and A=a*, respectively, and the effect of A on Y is linear such that if a=a*+1 then the ATE at any level of A represents the change in Y for a one-unit increase in values of A, regardless of choice of *a*. We seek to re-write the causal estimand in terms of quantities that can be estimated from observed data. For example, the true data generating mechanism is represented by DAG 1 (**Figure 1**, *A*→*M*→*Y*), and we had a single data source containing A and Y in which standard identifiability assumptions were satisfied (e.g., exchangeability, positivity), the ATE would be identified by *E*[*Y*|*A* = *a*] − *E*[*Y*|*A* = *a*^*^]. But in our motivating scenario, this cannot be directly estimated because there is no data set in which both Y and A are measured. However, here, since M_1_ (a continuous variable) is the only mediator between A and Y (and there is no direct path from A to Y) and it is measured in both the early- and late-life cohorts, we can rewrite *E*[*Y*|*A* = *a*] as:

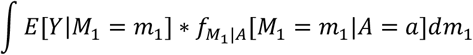

where *f* is the conditional density of M_1_ given A (note that because DAG 1 does not include a direct path from A to Y, Y is not conditioned on A). *E*[*Y*|*M*_1_] can be estimated in the late-life cohort (*i*.*e*., *E*[*Y*|*M*_1_, *R* = 1]) and 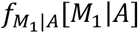 can be estimated in the early life cohort (*i*.*e*., 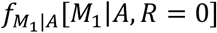). Because these are representative samples drawn from same target population, 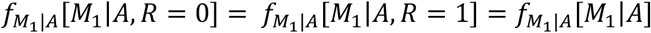, we can then estimate:

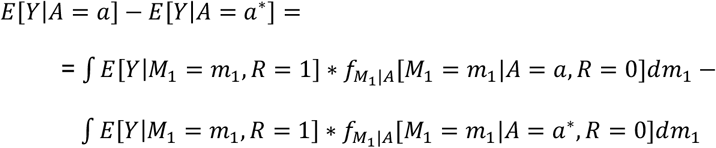

It is plausible to assume that the causal effect of A on Y is the same for the early- and later-life cohorts if, for example, both cohorts are randomly drawn from the same target population. However, if the effect of A on Y differs between cohorts (*e*.*g*., a selection node on Y), we may still identify the effect of A on Y in the late life cohort with weaker assumptions, specifically that the effect of A on M_1_ is the same for the two cohorts.

Above, if we were unable to assume the two cohorts were representative samples from the same target population, quantitative corrections such as weighting to account for recruitment bias or selective dropout, or transport methods such as weighting could be performed to estimate effects in other target populations.^32^ This requires additional assumptions and methods for transport, which have been previously well described.^22,28,30,34,40^ For simplicity, in this paper we assume both cohorts are a simple random sample of a shared target population to avoid the need for such corrections.

Because in our motivating question measures of A and Y exist in separate cohorts, the approach shown above fails if the data generating mechanism includes a direct (unmediated) effect from A to Y, such as in DAG 2 (*i*.*e*., we cannot estimate either *E*[*Y*|*M*_1_ = *m*_1_, *A* = *a, R* = 0] or *E*[*Y*|*M*_1_ = *m*_1_, *A* = *a, R* = 1]). Thus, in addition to the above sampling assumptions, and because A and Y are not both measured in any cohort, identifying the effect of A on Y in the synthetic cohort requires that A can be fully d-separated from Y in a given DAG. This requires that a set of variables that block all back *and* front door pathways between A and Y is measured in both cohorts. Intuitively, this is because we aim to create a synthetic cohort that reproduces all of the ways in which A and Y are associated in the target population. Thus, to capture those associations in the synthetic cohort, we need to combine datasets based on the variables that create them *i*.*e*., confounders and mediators along every back and front door pathway from A to Y. If A and Y remain d-connected given the matching set, the resulting combined data will either have residual confounding (due to an unblocked back door path) or the relationship between A and Y will be biased by mediating pathways that are unaccounted for in the synthetic cohort (due to an unblocked front door path). In practice, this necessitates measuring the merging variables at similar ages across the two cohorts. For example, for DAG 1, secular trends in the values of A, M_1_, or Y do not necessarily preclude this approach, but would raise doubts that the effect of A on M_1_ would be similar across the cohorts. Additionally, to identify the ATE in the synthetic cohort, positivity must be met in this merging step, specifically, for every stratum of merging variables in the later life (i.e., index) cohort the probability of observing that stratum must be non-zero and the probability of observing A within that stratum must be non-zero in the early life cohort (for DAG 1, *if P*(*M*_1_|*R* = 1) > 0 *then P*(*M*_1_|*R* = 0) > 0 *and P*(*A* = *a*|*M*_1_, *R* = 0) > 0 for all *a* ∈ *A*, although this could be generalized to other DAGs by considering M_1_ as a representing a vector all matching variables).

Finally, to estimate causal effects of A on Y in the synthetic cohort, we also require the typical assumptions for causal estimation – conditional exchangeability, positivity, correctly specified models, and the stable unit treatment value assumption (SUTVA) (detailed in **Appendix 1**)^41,42^ - to be fulfilled in the late life cohort. Additional assumptions sometimes included as part of “identifiability assumptions”^31,33^, *e*.*g*., consistency, temporality, no measurement error, are implicitly encoded in our DAGs.

#### (3) Overall simulation and matching approach

Two cohorts of size N=2,500 were randomly sampled from the data generating rules dictated by each DAG (simulation equations and parameter inputs for each DAG are listed in **Table 1**). In brief, we assigned values of continuous variables C, A, M_1_, M_2_, and Y by simulating each exogenous variable and then simulating each endogenous variable as a function of its parents. The true effect size for A on Y was 0.3 and the strength of confounding (*i*.*e*., the difference between the true effect and the estimated effect if the confounder is not adjusted for), when present in the DAG, was approximately equal to the true effect size, unless otherwise specified. In the early life cohort only A, M_1_, M_2_, and C were measured and in the later life cohort only M_1_, M_2_, C, and Y were measured. N=2,500 was chosen to reflect a reasonably large but realistic sample size for a cohort study in which likely important matching variables are available for the example relationship between early-life blood pressure and later dementia risk (*e*.*g*., anthropometric measures, blood biomarkers, genes).

After sampling each cohort, the synthetic cohort was created via a one-to-many matching procedure based on weighted Euclidean distance. The goal in creating the synthetic cohort is that every individual with an outcome in the late-life dataset is assigned the exposure values of their likely counterparts in the early-life cohort. The best early-life counterparts to provide this exposure information are those with the most similar values of variables included in the matching set. For example, if matching on M_1_ only, an observation in the late-life cohort for which M_1_=m_1_ was matched to multiple observations in the early-life cohort providing the best (closest) matches to M_1_=m_1_. In contrast, if matching on multiple mediators and/or mediators and confounders, best matches needed to be identified based on multiple variables (*e*.*g*., M_1_=m_1_, M_2_=m_2_, C=c). One-to-many matching was used to appropriately reflect the uncertainty in assigning individuals in the late-life cohort their exposure value based on the experiences of individuals in the early-life cohort.

Distances between matches were computed as follows. The distance between a given older-cohort individual and *every* individual in the early-life cohort was computed as the weighted Euclidean distance of (i.e., sum of squared differences in) the matching variables, with smaller numbers indicating a closer/better overall match. For each matching variable, distance was weighted by the inverse R^2^ value from a univariable model regressing A against that variable.

This was done so that the matching variables more strongly associated with A (that therefore improve our ability to replicate the data generating structure in the synthetic cohort) are given priority (*i*.*e*., upweighted) among all variables considered in the matching process. For each individual in the older cohort, we selected the 10 best matches from the early-life cohort. For comparison, we repeated all simulations selecting the 30 best matches instead of 10.

**Table 2** outlines the various simulation scenarios examined across all DAGs. For DAGs 1 and 2, all available matching variables were used to create the synthetic cohort. For other DAGs, the following variations on simulation procedures were examined:

1. For data generated under DAGs 3a-b and 4, we varied the availability of the variables used in the matching procedure to examine the consequences of incomplete d-separation. For DAG 3, the ATE is identifiable when M_1_ and M_2_ are both available for matching, so we examined scenarios where M_1_ was available but M_2_ was absent from both cohorts (scenario A) and where both were available for matching (scenario B). For DAG 4, we considered scenarios in which C was unmeasured in both cohorts (scenarios A,B,C) vs. available in both cohorts (scenarios D,E,F).
2. For data generated under DAG 4, we varied the strength of the relationship between the matching variables and the exposure, A, to examine the consequences of upweighting stronger predictors among the matching variables in the matching procedure. Specifically, we examined scenarios where C→A and A→M_1_ were of approximately equal strength (scenarios A & D), where C→A was stronger than A→M_1_ (scenarios B & E), and where C→A was weaker than A→M_1_ (scenarios C&F).

As a visual aid to linking the causal scenarios with the variables available in the observed data, in the rightmost columns of **Figure 1** the DAGs are redrawn and color-coded to show in which cohort variables are measured. Unless otherwise noted, we assume that participant age ranges in the cohorts are partially overlapping, and that the variables on which the cohorts are matched are measured at the same ages.

#### (4) Analyses

For DAGs 1-3, the ATE is identified in the synthetic cohort by the statistical parameter *E*[*Y*|*A* = *a*] − *E*[*Y*|*A* = *a*^*^]; for DAG 4, it is identified by *E*_*c*_{*E*[*Y*|*A* = *a, c*] − *E*[*Y*|*A* = *a*^*^, *c*]}. In the synthetic cohort, we estimated the ATE of A on Y and compared this to the true ATE of 0.30 (i.e., the ATE that would be estimated in the target population if all variables were measured on the same individuals) using OLS regression. To appropriately characterize uncertainty, we used a resampling with replacement such that each index case and each of its 10 (or 30) matches were randomly resampled into one of 10 (or 30) datasets and linear regression analyses were estimated in each. The estimated ATE was computed as the average across the datasets (*i*.*e*., matches) and the variance in the estimated ATE was computed using Rubin’s rules as the sum of within and across dataset (*i*.*e*., match) variance.^43^ The total variance was used to compute 95% confidence intervals (CI) for each estimated ATE in each dataset and then 95% CI coverage was computed as the proportion of datasets in which the 95% CI covered the true ATE value.

For each DAG, OLS regression models were of the following functional forms:

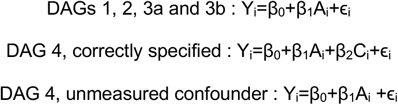

The coefficient of interest was β_1,_ the difference in Y associated with changing A from a* to a=a*+1 (a 1-unit increase). Note that in most scenarios these models are correctly specified: they use the same functional form that was used to generate the data in each DAG; however, in DAG 4, we additionally have a misspecified “unmeasured confounder” model for the scenarios where C was unmeasured and therefore unable to be matched on or adjusted for in analyses.

We evaluated estimation performance using the following set of performance metrics assessed across all 1,000 simulation runs: i) average matching-corrected standard error across estimates of the coefficient on A (SE); ii) bias of the estimated ATE calculated as the average difference between the coefficient for A in the model estimated in the synthetic cohort and the value of the coefficient on A in the model estimated in the target population; iii) mean square error (MSE) of the estimates of the coefficient on A (calculated as bias^2^ + variance across simulation estimates); and iv) mean 95% CI coverage. All simulations were performed using R 3.6.0.^44^ All code is publicly available on Github (*url blinded for review*).

## Results

As anticipated, in scenarios where complete d-separation by matching variables was possible, there was negligible bias in the estimated ATE in the synthetic cohort (**Table 2**). For example, in a simple setting with a causal sequence from A to Y through M_1_ (DAG 1), with matching on M_1_, the synthetic cohort performed well (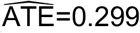, mean SE=0.032, mse=0.00026). In contrast, in DAG 2, where A influenced Y via a mechanism unrelated to the matching variable (i.e., there is a direct path from A to Y not via M_1_), the estimated ATE in the synthetic cohort was biased in a negative direction, as expected (bias=-0.080).

In DAGs 3a and 3b another linking variable, M_2_, was added along the direct path from A to Y. Consistent with DAGs 1 and 2, when this pathway through M_2_ was unblocked (i.e., when M_2_ was unmeasured and therefore unavailable for matching), the ATE estimate was biased. This bias was stronger in scenarios where M_2_ was an effect of the matching variable M_1_ (DAG 3a, bias=-0.140) rather than a cause of M_1_ (DAG 3b, bias=-0.053). In both DAGs, having a measure of M_2_ available for matching in addition to M_1_ nearly eliminated this bias.

In DAG 4, variable C confounds the relationship between A and Y (which are linked via M_1_). When C was unmeasured, and therefore unavailable for matching or adjustment in the synthetic cohort, the estimated ATE was biased in a positive direction. When the strength of relationships between C→A and A→M_1_ was approximately equal (scenario A), bias = 0.10; this bias was greater when C→A was stronger than A→M_1_ (scenario B, bias=0.209) than when A→M_1_ was stronger than C→A (scenario C, bias=0.027). In scenarios D-F, when C was available for matching and then subsequently adjusted for in analyses in the synthetic cohort, this bias was nearly eliminated.

Increasing the number of matches from 10 to 30 (**Appendix Table 1**) or using a weighted vs unweighted distance matching approach when matching on more than one covariate (**Appendix Table 2**) had minimal impact on bias and variance estimates. Across all simulation scenarios, simulation performance was excellent. For example, in scenarios where no bias was expected, bias was nearly 0 and MSE was low; in addition, although in these scenarios 95% CI coverage was generally higher than 0.95, this is unsurprising given we intentionally computed overly conservative variance estimates to appropriately characterize the uncertainty involved in our approach (i.e., matching different people from different datasets to create an individual life course history).

## Discussion

In this work, we consider scenarios in which two partially age-overlapping studies covering different life course periods are combined to create a synthetic cohort with longitudinal data from early to later life. We used simulations to examine a novel, flexible matching-based approach to creating this life course synthetic cohort when exposure and outcome information are not available in the same component studies. Our simulations considered a range of possible causal structures to establish the feasibility of evaluating effects of early life risk factors on late life outcomes in the synthetic cohort. Using the shared variables as a basis for matching, we were able to recover the ATE from the target population using the synthetic cohort in many but not all causal scenarios. Scenarios in which the matching variables d-separate the exposure and outcome can deliver unbiased effect estimates. When A and Y remained d-connected given the matching set, there was either residual confounding due to an unblocked back door path (e.g., DAG 4 scenarios A-C) or bias due to mediating pathways that were not accounted for (i.e., due to an unblocked front door path e.g., DAG 2).

By clarifying the assumptions necessary for unbiased effect estimation in the synthetic cohort, we illuminate the challenges investigators will need to overcome when pooling different longitudinal cohorts in practice. A critical assumption of our approach is that it is conceptually reasonable to think of the two cohorts as drawn from either the same population or from two populations with the same causal structure. Although this may rarely be true, there may be many situations in which it is acceptable to assume that the two populations share a causal structure with respect to the variables of interest. However, this remains a major simplifying assumption in our simulations – specifically, by considering the earlier and later-life cohorts as simple random samples drawn from the same underlying target population at different ages, we essentially assume the older cohort represents later life experience of the younger cohort had we been able to continuously follow them over time, creating a situation in which the relationship of interest (e.g., A→M_1_, M_1_→Y) would be the same in each cohort. There are several ways in which this assumption is likely violated in practice, including differential participation or survival processes operating in each cohort. Period effects or secular trends in the relationships that differ between cohorts may also violate the assumption. The performance of our approach when these types of violations are encountered is an important aspect of future work in this area.

Another assumption of our approach, which in practice may be more challenging, is that the matching variables adequately capture all backdoor and frontdoor pathways between the exposure and the outcome (i.e., d-separate the exposure and outcome). Conceptually similar assumptions are implicit in much of epidemiology (e.g., no unmeasured confounding). In reality, while *complete* d-separation of exposure and outcome is likely not possible, it is conceivable that major mediators and/or confounders of any early-life exposure and later-life outcome, such as core sociodemographic variables and/or general health measures, are common across multiple cohorts, albeit potentially measured in different ways. These shared variables would provide the basis for the linking and matching process that we propose. Further, in real data sets tools like e-values and/or bounds could help gauge the sensitivity of synthetic cohort estimates to incomplete d-separation by providing a range of plausible values for the ATE under varying assumptions about the proportion of confounding and/or mediating pathways *not* captured in the matching procedure. Future work on pooling studies to create synthetic life course cohorts would benefit from exploring these tools for issues of incomplete d-separation and measurement error. Finally, our approach will never outperform results that could be achieved if all variables were measured in the full target population. Therefore, all of the conventional assumptions for causal inference must hold (e.g., exchangeability, positivity).

Although simplifying assumptions are common in simulation studies, our findings must be interpreted in light of these simplifications. For one, we explored only a limited number of causal settings, we only considered pooling of two cohorts, and the only estimand considered was the ATE. Future simulation work expanding on this project should consider other perhaps more complicated causal structures, allow for combination of more than two data sources, and examine the performance of alternative estimands in the synthetic cohort (e.g., direct effects, which may have different patterns of bias across causal structures). Additionally, the computational time increases as the number of matching variables increase, and this could be an important limitation of our approach in practice, when many more variables need to be included in the matching set. Another simplification in our simulations was the assumption that all variables were measured without error. Thus, additional future directions for this work include developing tools to address measurement error in the matching variables, use of slightly different measurement instruments so matching can be based on latent variables rather than observed variables, and applied research on the most relevant variables to include for matching for specific exposure-outcome relationships of interest. Finally, we only examined one distance matching-based approach to combining cohorts in our simulations. Matching is a flexible approach that allows for strong covariate control without inter/extrapolation if good matches can be found. However, as the number of matching variables grows, the availability of good matches may decrease, raising concerns about practical positivity violations; in this situation, it is possible that other data pooling approaches (e.g., imputation-based) may outperform matching. Thus, it would be important for future work to compare the performance of different matching-based approaches, as well as compare matching-based approaches with other more commonly used approaches to pooling life course data from different cohorts, such as model-based imputation.

## Conclusions

Evaluating early- and mid-life determinants of late-life disease is a high priority in health and life course research.^45^ To help concretize our simulation, we used an example from dementia research to demonstrate the types of exposure, merging, and outcome variables that may be of interest (e.g., early and mid-life blood pressure and dementia risk score), but our approach is broadly applicable to a wide range of life course research questions (e.g., early life exposure to pollution and later-life lung function or cancer risk). Life course research is often stymied by the limited follow-up or lack of detailed early life measures available in cohorts of older adults. The matched cohort approach we propose affords opportunities to evaluate life course risk factor effects on late life outcomes.

## Data Availability

The data is simulated and thus data sharing is not applicable.

---

**Appendix Table 1.** Average estimates in synthetic cohort (N=2,500) created using 1:30 matching^1^ across 1,000 simulation iterations

| DAG | Scenario | Y data generation | Strength of matching variables with A | Variables measured in young cohort | Variables measured in old cohort | Matching variable(s) | Linear regression model in synthetic cohort | ATE in synthetic cohort | SE<sup>2</sup> | Bias<sup>3</sup> | MSE | 95% CI coverage |
| --- | --- | --- | --- | --- | --- | --- | --- | --- | --- | --- | --- | --- |
| 1 | - | Y~M<sub>1</sub> | C→A ≅ A→M<sub>1</sub> | V, M<sub>1</sub> | M<sub>1</sub>, Y | M<sub>1</sub> | Y=A | 0.297 | 0.031 | -0.003 | 0.0002 | 0.986 |
| 2 | - | Y~A+M<sub>1</sub> | C→A ≅ A→M<sub>1</sub> | V, M<sub>1</sub> | M<sub>1</sub>, Y | M<sub>1</sub> | Y=A | 0.218 | 0.030 | -0.082 | 0.0069 | 0.145 |
| 3a | A | Y~M<sub>1</sub>+M<sub>2</sub> | C→A ≅ A→M<sub>1</sub> | V, M<sub>1</sub> | M<sub>1</sub>, Y | M<sub>1</sub> | Y=A | 0.158 | 0.034 | -0.142 | 0.0203 | 0.005 |
| 3a | B | Y~M<sub>1</sub>+M<sub>2</sub> | C→A ≅ A→M<sub>1</sub> | V, M<sub>1</sub>, M<sub>2</sub> | M<sub>1</sub>, M<sub>2</sub>, Y | M<sub>1</sub>, M<sub>2</sub> | Y=A | 0.293 | 0.032 | -0.007 | 0.0003 | 0.989 |
| 3b | A | Y~M<sub>1</sub>+M<sub>2</sub> | C→A ≅ A→M<sub>1</sub> | V, M<sub>1</sub> | M<sub>1</sub>, Y | M<sub>1</sub> | Y=A | 0.247 | 0.032 | -0.053 | 0.0030 | 0.640 |
| 3b | B | Y~M<sub>1</sub>+M<sub>2</sub> | C→A ≅ A→M<sub>1</sub> | V, M<sub>1</sub>, M<sub>2</sub> | M<sub>1</sub>, M<sub>2</sub>, Y | M<sub>1</sub>, M<sub>2</sub> | Y=A | 0.292 | 0.032 | -0.008 | 0.0003 | 0.976 |
| 4 | A | Y~M<sub>1</sub>+C | C→A ≅ A→M<sub>1</sub> | V, M<sub>1</sub> | M<sub>1</sub>, Y | M<sub>1</sub> | Y=A | 0.398 | 0.029 | 0.098 | 0.0099 | 0.048 |
| 4 | B | Y~M<sub>1</sub>+C | C→A > A→M<sub>1</sub> | V, M<sub>1</sub> | M<sub>1</sub>, Y | M<sub>1</sub> | Y=A | 0.508 | 0.028 | 0.208 | 0.0436 | 0 |
| 4 | C | Y~M<sub>1</sub>+C | C→A < A→M<sub>1</sub> | V, M<sub>1</sub> | M<sub>1</sub>, Y | M<sub>1</sub> | Y=A | 0.325 | 0.034 | 0.025 | 0.0009 | 0.938 |
| 4 | D | Y~M<sub>1</sub>+C | C→A ≅ A→M<sub>1</sub> | V, M<sub>1</sub>, C | C, M<sub>1</sub>, Y | M<sub>1</sub>, C | Y=A+C | 0.291 | 0.030 | -0.009 | 0.0003 | 0.975 |
| 4 | E | Y~M<sub>1</sub>+C | C→A > A→M<sub>1</sub> | V, M<sub>1</sub>, C | C, M<sub>1</sub>, Y | M<sub>1</sub>, C | Y=A+C | 0.291 | 0.029 | -0.009 | 0.0003 | 0.973 |
| 4 | F | Y~M<sub>1</sub>+C | C→A < A→M<sub>1</sub> | V, M<sub>1</sub>, C | C, M<sub>1</sub>, Y | M<sub>1</sub>, C | Y=A+C | 0.291 | 0.030 | -0.027 | 0.0003 | 0.978 |

<sup>1</sup>Matching approach used was based on weighted Euclidean distance where weights were the inverse R<sup>2</sup> value from a univariable model regressing A. This was done so that the matching variables more strongly associated with A were upweighted among all variables considered in the matching process.
<sup>2</sup>Calculated using Rubin's rules as the square root of total within- and between-match variance
<sup>3</sup>Calculated as average $\overline{ATE}$ in synthetic cohort – ATE in target population
<sup>4</sup>MSE calculated as bias<sup>2</sup>+variance
<sup>5</sup>Mean of the 95% confidence interval coverages across simulation iterations; within simulation iterations, 95% CI coverage is the proportion of the 95% CI in each of the 10 datasets (i.e., matches) that covered the true value of 0.30.

**Appendix Table 2.**
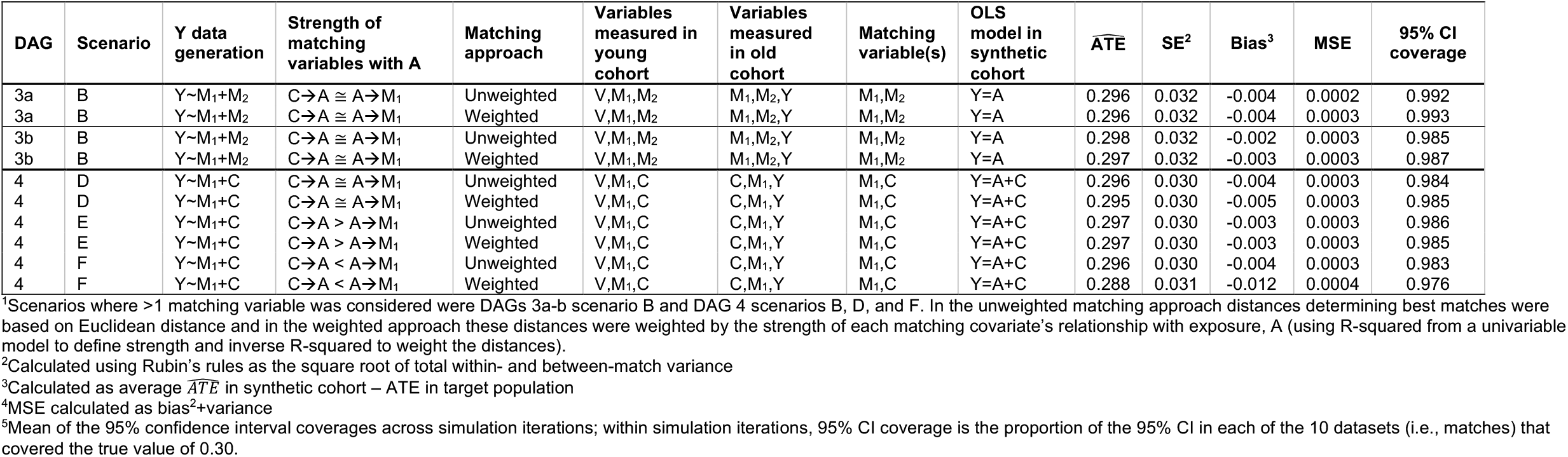
Average estimates in synthetic cohort (N=2,500) created using 1:10 matching across 1,000 simulation iterations, comparing unweighted vs. weighted distance matching approach when matching on >1 covariate^1^.

## Appendix 1

Assuming the early- and late-life cohorts are simple random samples from the same population and the set of matching variables used to link observations in the early and late-life cohorts fully d-separate A and Y, then the ATE can be identified in the synthetic cohort if the following identifiability assumptions are met. Note that these are the same assumptions that would be required if we conducted the analysis with measurement of all variables in the target population:

1. <u>Exchangeability</u>: The potential outcomes for Y under any potential value of A (*Y*_*a*_) are independent of received value of A conditional on the covariate set C: *Y*_*a*_ Џ *A* | *C* for all values of A. For our causal scenarios, if any backdoor pathways d-connect A and Y in the target population, then merging on the variables that block these pathways (e.g., C) reproduces them in the synthetic cohort, biasing estimates of the ATE if unaccounted for. To satisfy conditional exchangeability, these backdoor paths must be additionally dealt with in the analysis conducted within the synthetic cohort, for example by conditioning on the variables that block them.
2. <u>Positivity</u>: In general, for a discrete exposure A and a vector of covariates, Z, the probability of A=a is nonzero within every level of observed Z (or *P(A = a* | *z) > 0* for all *a* ∈ *A*). For a continuous exposure, positivity can be expressed as the density of A=a is greater than zero within every level of observed Z (or *f*(*A* = *a*|*Z*) > 0 for *all a* ∈ *A*).
3. <u>SUTVA</u>: We assume that the effect of the exposure on the outcome for an individual does not depend on the exposure values of other individuals, for example there can be no “interference” or herd effects.^36,37^
4. No model misspecification: where parametric models are used for inference in the synthetic cohort, we assume no model misspecification.

